# Results of an early second PCR test performed on SARS-CoV-2 positive patients may indicate risk for severe COVID-19

**DOI:** 10.1101/2021.02.09.21251371

**Authors:** Barak Mizrahi, Maytal Bivas-Benita, Nir Kalkstein, Pinchas Akiva, Chen Yanover, Yoav Yehezkelli, Yoav Kessler, Sharon Hermoni Alon, Eitan Rubin, Gabriel Chodick

## Abstract

Identifying patients at increased risk for severe COVID-19 is of high priority during the pandemic as it could affect clinical management and shape public health guidelines. In this study we assessed whether a second PCR test conducted 2-7 days after a SARS-CoV-2 positive test could identify patients at risk for severe illness. Analysis of a nationwide electronic health records data of 1,683 SARS-CoV-2 positive individuals indicated that a second negative PCR test result was associated with lower risk for severe illness compared to a positive result. This association was seen across different age groups and clinical settings. More importantly, it was not limited to recovering patients but also observed in patients who still had evidence of COVID-19 as determined by a subsequent positive PCR test. Our study suggests that an early second PCR test may be used as an additional risk-assessment tool to improve disease management and patient care.

## Introduction

The COVID-19 pandemic is continuing to spread, surpassing 92M confirmed cases and 2M deaths globally as of January 2021 ^1^. Although survival rates have gradually improved by the development of successful treatment protocols for moderate and severe patients, the disease continues to claim lives and containment is proven difficult ^2–6^. Widespread severe acute respiratory syndrome coronavirus 2 (SARS-CoV-2) testing is regarded as best practice for detection of infection and epidemiological surveillance. Strategies for detection and containment of COVID-19 have evolved and now that testing has become more available it is performed both in community-based sites and in hospitals. Multiple testing following exposure and diagnosis of COVID-19 patients has been used to evaluate infectiousness and assure resolution of infection, especially considering the known silent spread by infected individuals that are asymptomatic ^7–11^.

Current SARS-CoV-2 infection is mostly tested using reverse transcription quantitative polymerase chain reaction of viral genes (RT-qPCR)-based assays, determining presence of viral genetic signatures ^12^. This test is highly specific, resulting in minimal false-positive rates and is considered the most accurate COVID-19 diagnosis method ^13,14^. However, a positive PCR test is unable to report the exact disease timeline of the patients, detect whether they are still infectious or determine what will be the severity of their disease. It was therefore suggested that frequent testing serve as a surveillance testing regimen to enable better detection of the infectious window and disease course ^15^.

While multiple testing in short time intervals may complement the current diagnostic tests and draw a more complete clinical timeline of SARS-CoV-2 infected individuals, it is important to realize the considerable rates of false-negative results in SARS-CoV-2 positive patients ^16^. False-negative results can be due to technical errors during sample collection or assay execution but may also appear due to low viral load resulting in RNA levels under the detection limit ^17^. Other reports showed that COVID-19 recovered patients (who tested positive and then presumed recovered by subsequent negative tests) could be SARS-CoV-2 positive again during their isolation period. This highlighted that negative test results can appear during the disease timeline but not necessarily determine infection resolution ^18^. While multiple testing may unfold the disease timeline, it is unclear whether it could indicate disease severity following COVID-19 diagnosis.

To evaluate whether additional testing following diagnosis could indicate severe illness, we analyzed electronic health records (EHR) from Maccabi Healthcare Services (MHS), the second largest Health Maintenance Organization (HMO) in Israel, which included longitudinal results of SARS-CoV-2 testing during the COVID-19 outbreak. Our study investigated the association between PCR results of a second test performed 2-7 days after a positive COVID-19 diagnosis and deterioration to severe disease. We analyzed different age groups, different clinical settings and patients at different stages in their disease timeline with the aim of supporting physicians’ risk assessment early for providing appropriate care to those at risk.

## Results

Data from March 1, 2020 to August 9, 2020 included 239,048 adults with records of a SARS-CoV-2 PCR test, among them 15,822 (6.6%) tested positive for the virus (Table 1). Tested individuals had an average age of 44.1 years and males were 52% of those tested positive. Among those tested positive, we identified 1,683 individuals who also had a second PCR test performed 2-7 days following diagnosis and their disease outcomes were analyzed throughout our study. Another subpopulation we explored was of 3,135 adults who had additional multiple PCR tests between their first and last documented positive SARS-CoV-2 test (defined as a SARS-CoV-2 positive window; see in “Methods” section).

**Table 1.** Baseline characteristics of study cohort of SARS-CoV-2 tested individuals.

| Characteristic | All individuals<br>n=239,048<br>(100%) | COVID-19<br>negative<br>n=223,226<br>(93.4%) | COVID-19<br>positive<br>n= 15,822<br>(6.6%) | COVID-19 positive with tests<br>within the SARS CoV-2<br>“positive window”<br>n=3,135 |
| --- | --- | --- | --- | --- |
| Age, years ± SD | 44.1 ± 18.6 | 44.3 ± 18.7 | 40.6 ± 17.1 | 43.3 ± 18.7 |
| Number of males<br>(% of population) | 103,231 (43%) | 94,933 (43%) | 8,298 (52%) | 1,719 (55%) |
| Average number<br>of tests per<br>individual | 1.57 ± 1.34 | 1.47 ± 1.19 | 3.00 ± 2.29 | 6.07 ± 2.44 |
| Total number of<br>tests | 375,558 | 328,159 | 47,399 | 19,025 |
| Number of tests after infection |  |  | 27,796 | 15,289 |

\* "Positive window" refers to the COVID-19 positive period in individuals who had additional PCR tests between their first and last documented positive SARS-CoV-2 tests.

The probability to develop a severe disease was evaluated in the study’s population stratified to three age groups: 18-59, 60-79 and ≥80 (Table 2). The overall probability to develop severe COVID-19 was 2.63% in the adult population. The probability of the 18-59 years old age group to deteriorate to severe condition was 0.99% (95% confidence interval (CI) = 0.83-1.16%). The high-risk population of adults older than 60 years showed increased probability to develop severe COVID-19 with a substantial difference between the age groups 60-79 and ≥80 years old (Probability=8.15% (6.98-9.32%) and 30.62% (26.69-34.54%), respectively).

**Table 2:** Age stratification of risk for deterioration in SARS-CoV-2-positive patients.

| Age (yrs) | # of patients with positive test | # of patients with severe condition | Probability to develop severe condition | 95% CI |
| --- | --- | --- | --- | --- |
| 18-59 | 13,478 | 134 | 0.99% | [0.83%-1.16%] |
| 60-79 | 1,939 | 158 | 8.15% | [6.98%-9.32%] |
| 80-100 | 405 | 124 | 30.62% | [26.69%-34.54%] |
| All | 15,822 | 416 | 2.63% | [2.38%-2.88%] |

### Clinical implications of a second testing following a SARS-CoV-2 positive PCR test

To evaluate whether an early second PCR test is indicative of clinical outcomes of COVID-19 we analyzed the clinical outcomes of SARS-CoV-2 positive individuals who had a second PCR test done within a week from the first positive test. Our analysis showed that a negative test result in the second test was correlated with a lower probability to deteriorate to severe COVID-19 in all age groups compared to a positive test (Fig. 1a; full results are presented in Supplementary Table 1). In the ≥80 years age group, a second positive test presented the highest probability for deterioration followed by the 60-79 and the 18-59 years age groups (41.76% (31.63-51.89%), 19.62% (13.43-25.81%) and 3.53% (2.08-4.97%), respectively). We also measured the probabilities for severe illness among individuals who were not tested a second time to assess if a bias towards a more severe outcome existed in those who tested. The probabilities for severe illness in patients not-tested a second time followed the same trend as those who tested (29.45% (24.07-34.84%) in the ≥80 years age group; 7.23% (5.99-8.47%) in the 60-79 years and 0.87% (0.70-1.03%) in the 18-59 years).

**Fig. 1.**
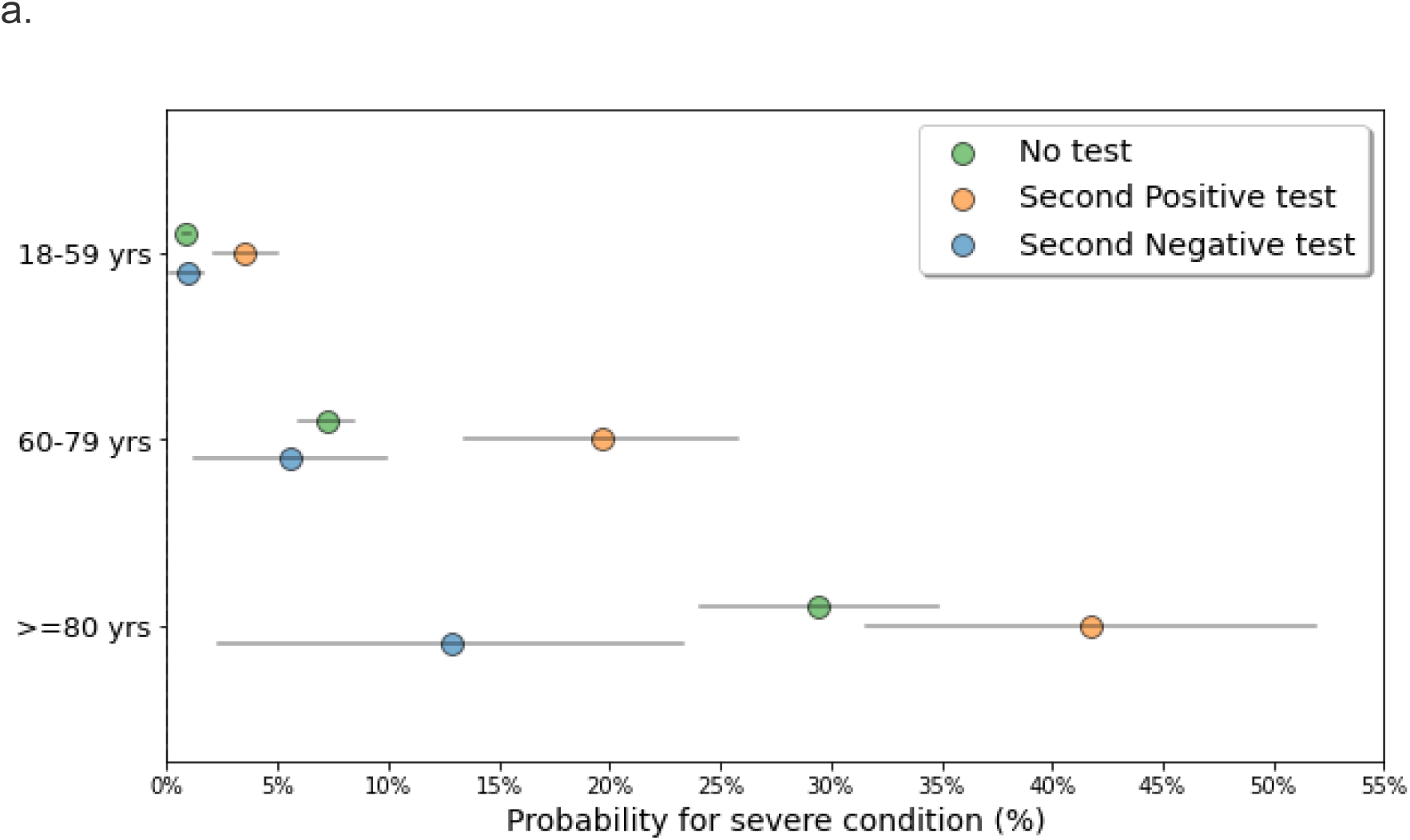
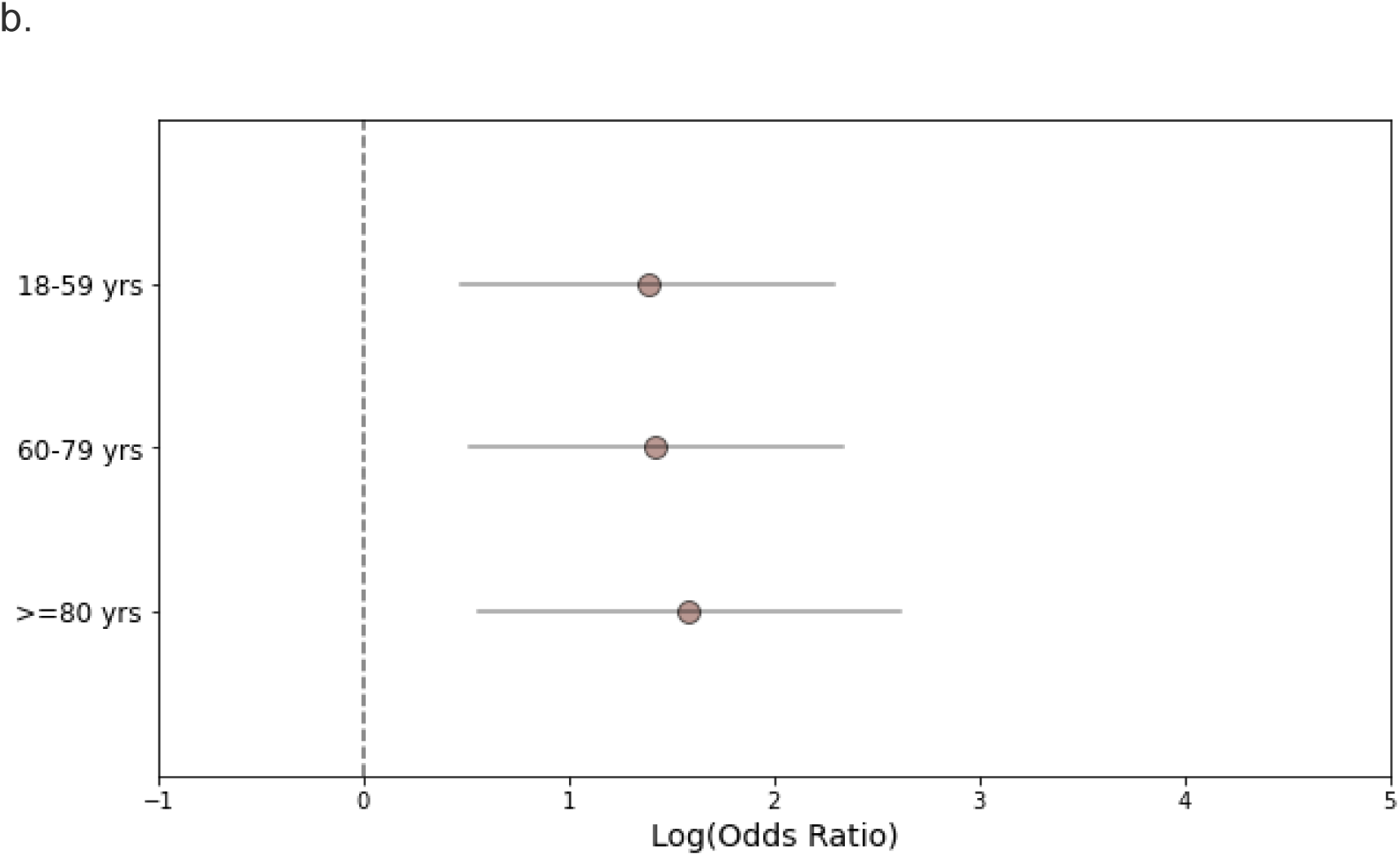
Deterioration probability and odds ratio analysis for SARS-CoV-2 positive individuals following a second PCR test. (a): Probability for severe COVID-19 in infected individuals tested a second time within the first week following diagnosis. Blue circles represent individuals who tested negative, orange circles represent individuals who tested positive and green circles represent individuals who were not tested in the week following diagnosis. Grey lines indicate 95% confidence intervals. (b): Log odds ratio (OR) for severe COVID-19 was calculated by the result of a second PCR test taken a week following diagnosis in a defined age group (18-59 years, n=1287; 60-79 years, n=266; >=80 years, n=130). OR>0 indicated larger risk in the population that tested positive compared to the population tested negative. Calculated log odds ratios are presented along with gray lines indicating 95% confidence intervals.

**Fig. 2.**
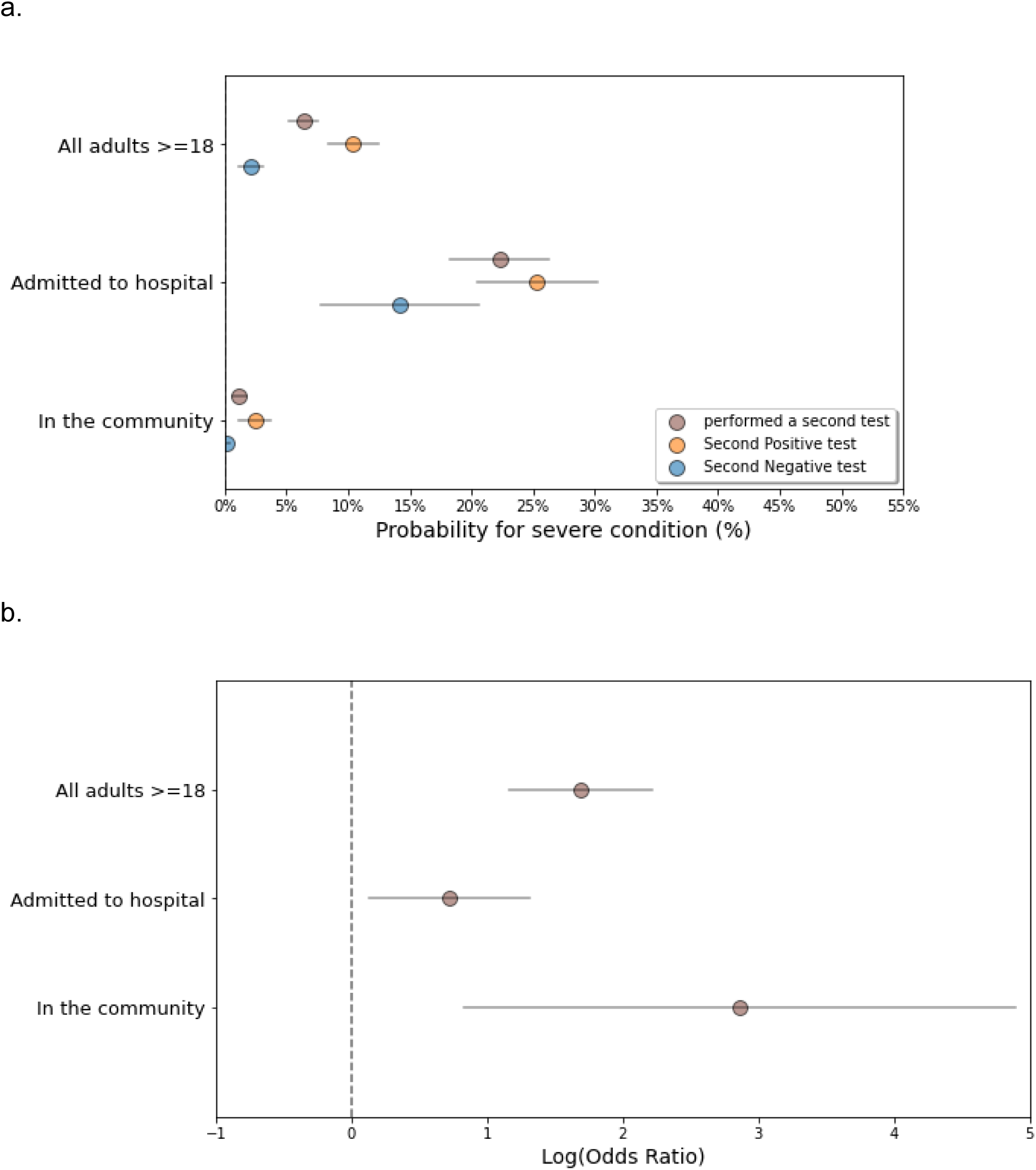
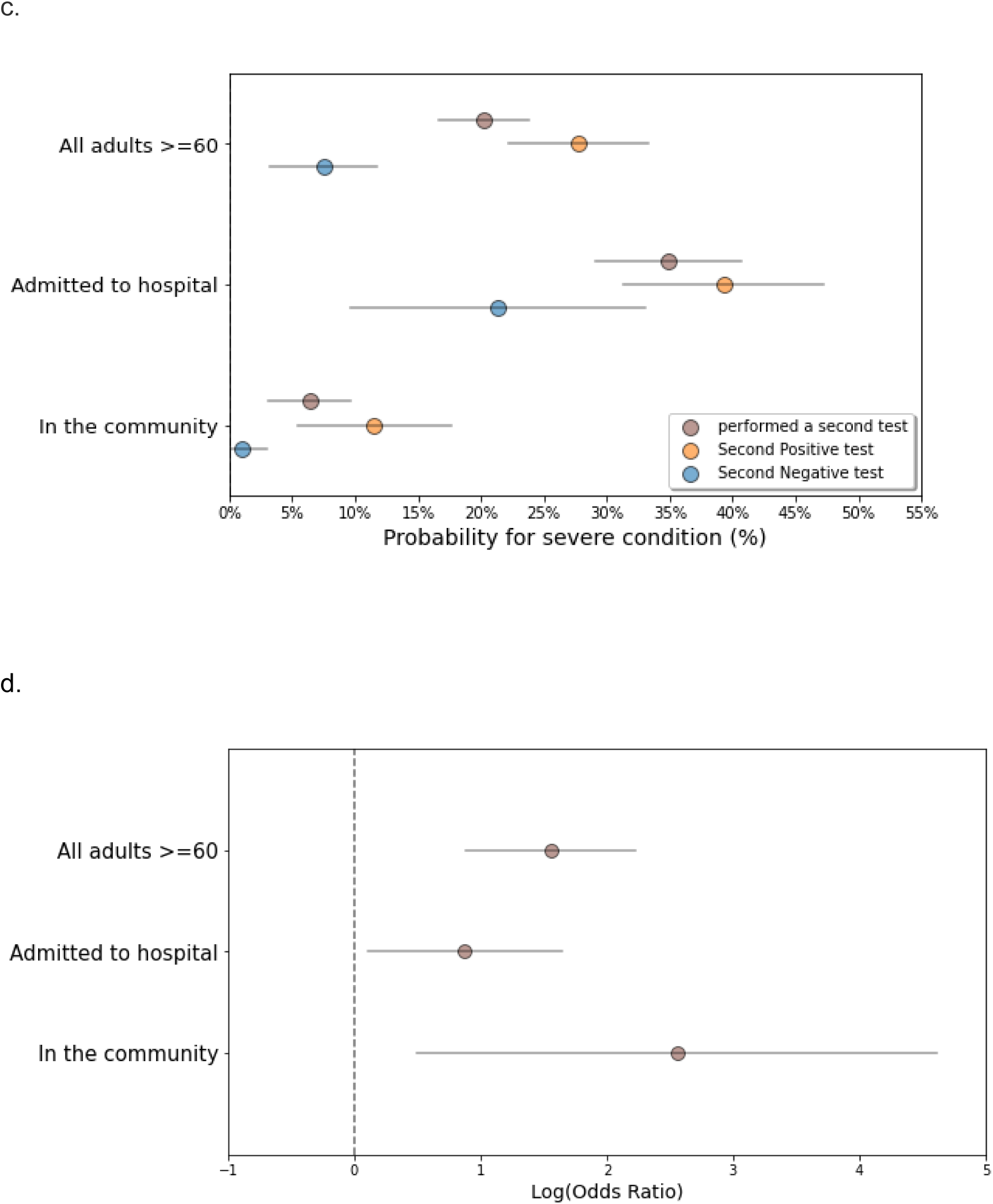
Probability analysis for COVID-19 deterioration following a second SARS-CoV-2 PCR test in different settings. (a): Probability of infected adults, tested a second time within the first week following diagnosis in the hospital and in the community, to deteriorate to severe COVID-19. Brown circles represent individuals with a second test a week following diagnosis, blue circles represent individuals who tested negative, orange circles represent individuals who tested positive. Grey lines indicate 95% confidence intervals. (b): Log odds ratio (OR) for severe COVID-19 was calculated by the result of a second PCR test taken a week following diagnosis in the hospital and in the community. OR>0 indicated larger risk in the population that tested positive compared to the population tested negative. Calculated log odds ratios are presented along with gray lines indicating 95% confidence intervals. (c): Probability analysis for individuals ≥60 years old. (d): Log odds ratio (OR) for severe COVID-19 for individuals ≥60 years old.

Despite the differences in the probability rates for deterioration in each age group, a positive test was highly associated with deterioration to severe COVID-19 compared to negative test across all ages (Fig. 1b; Supplementary Table 2). Odds ratio analysis in adults ≥ 80 years (odds ratio (OR) = 4.88 (1.75–13.62)), in 60-79 years (OR = 4.15 (1.67–10.33) and in 18-59 years (OR = 4.00 (1.61–9.94)) demonstrated the ability of the second early PCR test to distinguish between individuals at higher risk for clinical deterioration and those who will experience a milder disease.

### A negative SARS-CoV-2 PCR test result following COVID-19 diagnosis is associated with better outcomes in both hospital and community settings

Next, we investigated whether the association of the second test result with disease deterioration was dependent on the testing settings. We therefore sub grouped the SARS-CoV-2-positive patients to those tested the second time while admitted to the hospital vs those tested in the community and measured their outcomes. Furthermore, following the previous observations that the population ≥ 60 years is at an increased risk for severe disease and a second PCR test could differentiate between those who will deteriorate, additional analysis on this age subgroup was included (combining the data of both 60-79 and ≥ 80 years old populations). Results showed that 24.8% of adults ≥ 18 and 48.5% of ≥ 60 years old tested a second time within a week following diagnosis while they were admitted to the hospital (Supplementary Table 3). The risk for deterioration to severe illness in hospitalized patients was higher than in patients tested in the community and the older population was at increased risk in both settings (For all adults ≥ 18 years: Probability in the hospital = 22.3% (18.31%-26.30%) vs Probability in the community = 1.18% (0.59%-1.78%); For ≥ 60 years: Probability in the hospital = 34.9% (29.09%-40.70%) vs Probability in the community = 6.37% (3.12%-9.62%)). However, the result of the second SARS-CoV-2 PCR test was indicative for severe disease outcome in both settings with higher odds ratio in the community (OR=17.56 (2.30-133.93) for all adults; OR = 12.92 (1.64–101.2) for ≥ 60 years; Supplementary Table 4) compared to the hospital (OR=2.06 (1.14-3.70) for all adults; OR = 2.4 (1.11– 5.20) for ≥ 60 years). These observations highlighted that the second early PCR test result had clinical implications in COVID-19 diagnosed individuals, in both hospital and community settings, and that a negative test result was associated with lower probability for clinical deterioration.

### An early second negative PCR test in patients that did not recover, as determined by a successive positive test, is indicative of disease timeline and associated with better disease outcomes

Following confirmation of infection by a positive PCR test result, a “positive window” was detected in some patients, which is defined as the period between their first and their last positive test (from diagnosis date to the last positive test). In some patients additional PCR tests were performed within a “positive window” and when the results were negative they were mostly regarded as “false-negative” as they did not indicate disease resolution. To evaluate whether negative test results were still associated with better clinical outcomes, despite knowing viral RNA is still present, we analyzed data of adult patients with a “positive window” that included additional tests in it. A total of 5,823 tests from 3,094 patients with a positive window of up to 3 weeks following diagnosis was followed, of those, 1,799 (30%) were negative (Fig. 3; Supplementary Table 5). Higher negative test rates were observed the farther they were taken from the date of diagnosis, starting with 4.3% (0.6%-7.9%) on the first day from diagnosis and reaching 42.0% (35.0%-48.9%) by day 21. This increase in negative rate is seen despite being in a positive window and is affected by the time from diagnosis.

**Fig. 3.**
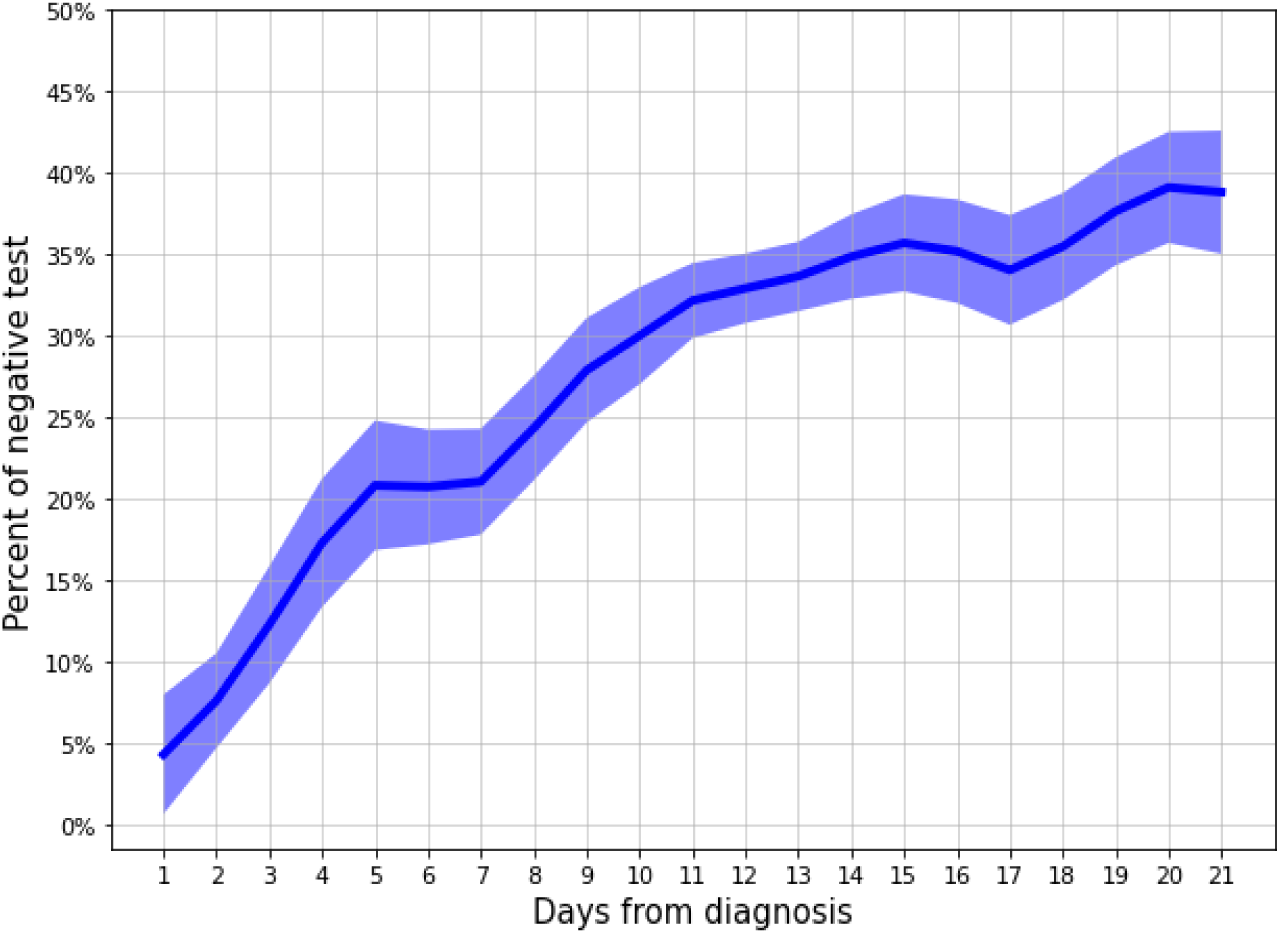
Negative rate of SARS-CoV-2 tests following a positive COVID-19 diagnosis within a positive window. Prevalence of negative test results in the population of adult individuals with a positive COVID-19 window, defined between the first and last positive test results taken. Line represents the negative tests rate from day 1 to day 21 following a positive COVID-19 diagnosis. Each time point is calculated by taking a 3 days window (±1 days from day measured) except in the first day which was taken as is. Shaded area represents 95% binomial proportion confidence intervals.

Negative COVID-19 test results within a “positive window” are considered insignificant as they do not reflect recovery from COVID-19. To evaluate whether an early second test was indicative of disease severity even in patients which we retrospectively knew had additional SARS-CoV-2-positive test, we analyzed 687 patients with a “positive window” and a second early PCR test (Fig. 4; Supplementary Table 6). We analyzed all patients who tested positive and divided them to those with a “positive window” and those without for comparison. Data showed that an early second negative test result in patients with a “positive window” was associated with decreased probability for severe illness compared to a positive test result (≥18 years (Fig. 4a): 3.2% (1.2-6.3%) vs 10.5% (8.0-13.0%); ≥ 60 years (Fig. 4c): 8.3% (0-19.4%) vs 26.6% (19.7-33.5%)). The association of a second negative test with decreased probability for severe illness was also seen in patients without the “positive window”. In the adults ≥18 years age group, the risk for disease deterioration in the individuals without an early second test was quite similar to that of those who tested negative, while in the ≥ 60 years age group it was slightly higher as it includes those who will become severe but have not tested a second time. An early second SARS-CoV-2 PCR test result was indicative of severe disease outcome in patients with a “positive window” and in patients without it in both age groups, presenting similar OR values (Fig. 4b and 4d; Supplementary Table 7). These results show that a second negative test in SARS-CoV-2 positive individuals is indicative for decreased probability of disease deterioration whether the patient is in a “positive window” or not.

**Fig. 4.**
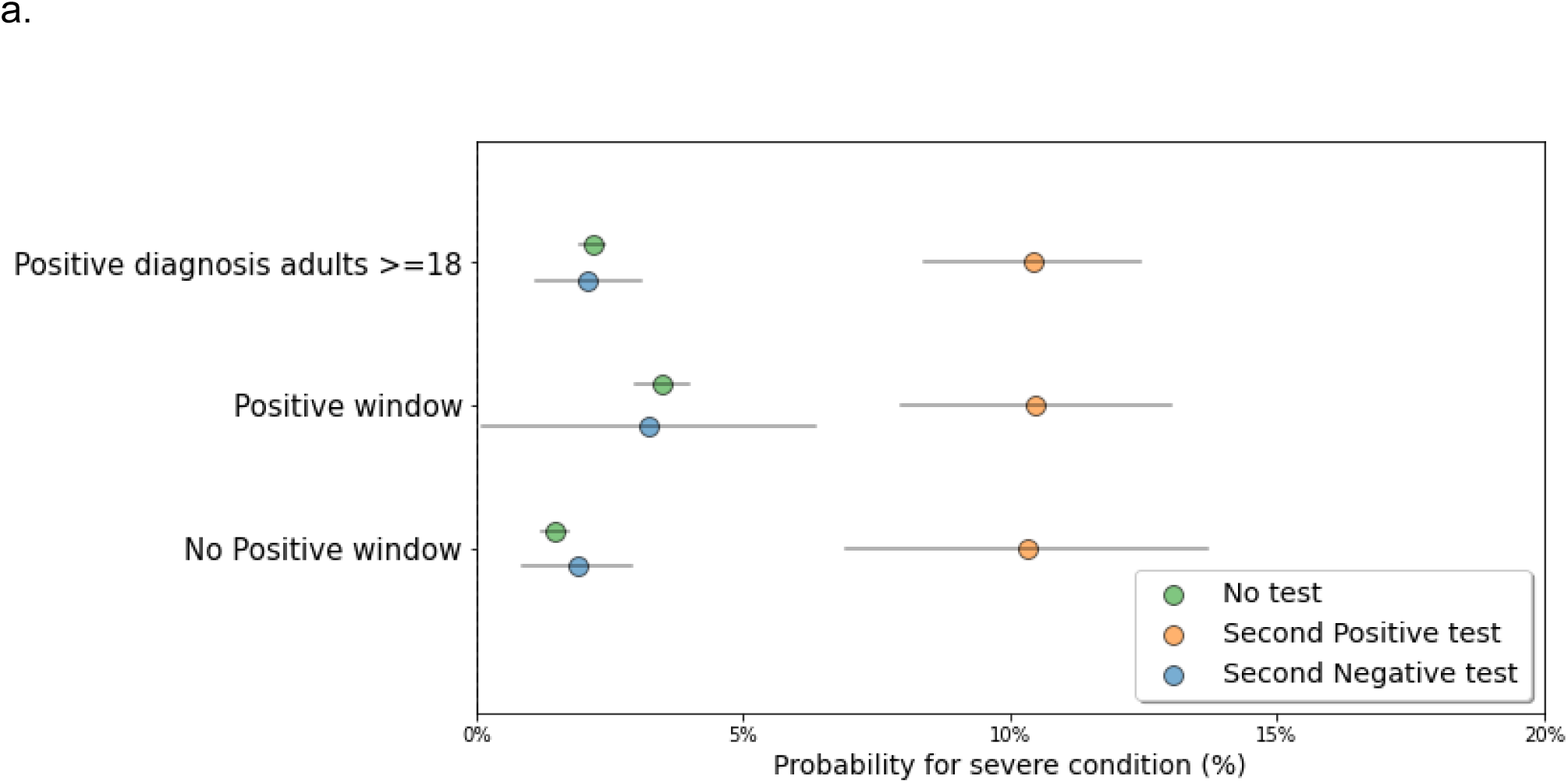
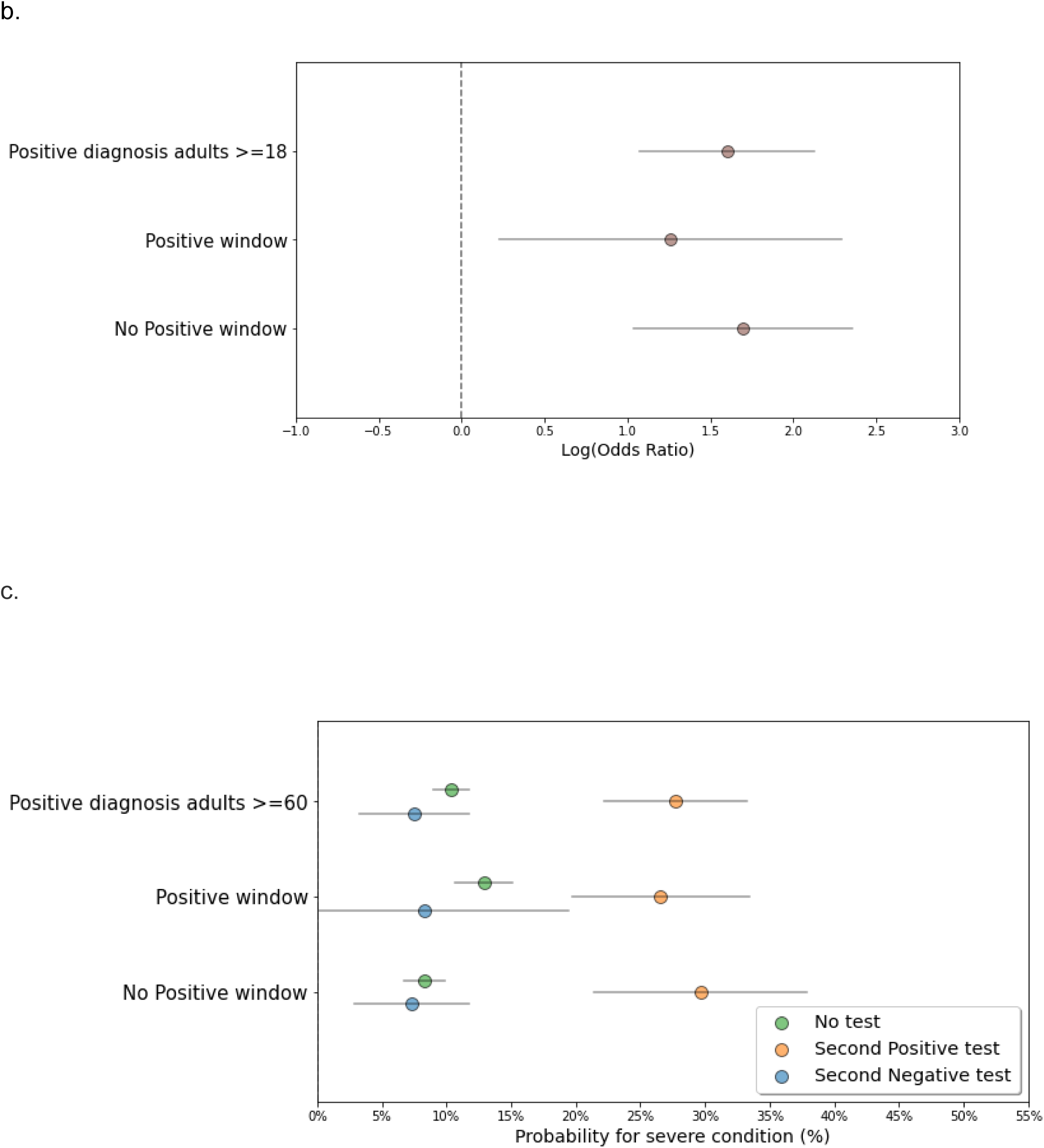
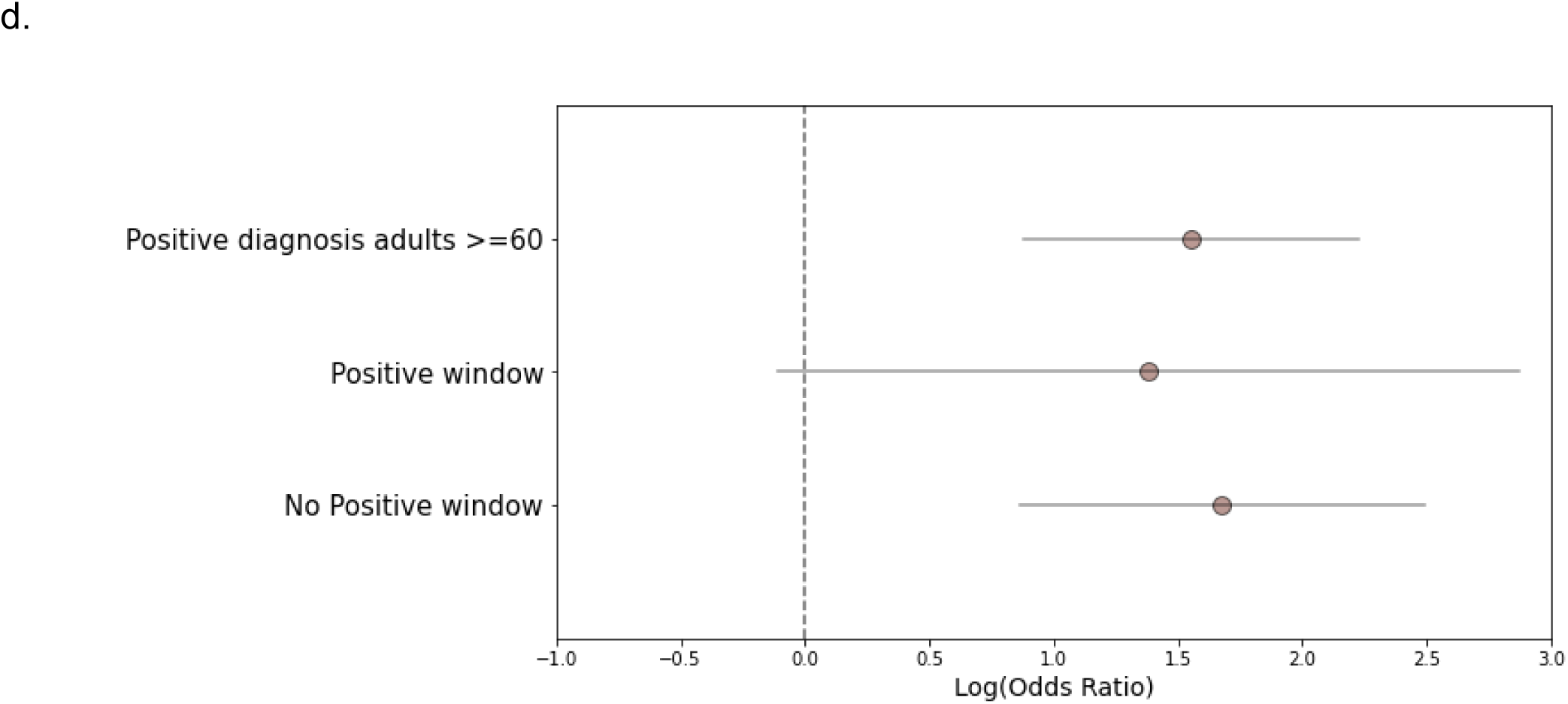
Probability analysis for disease deterioration following an early second SARS-CoV-2 PCR test in COVID-19 infected individuals with a “positive window”. (a): Probability of infected adults ≥18 years with a “positive window” tested a second time within the first week following diagnosis to deteriorate to severe COVID-19. Blue circles represent individuals who tested negative, orange circles represent individuals who tested positive and green circles represent individuals who were not tested in the week following diagnosis. Grey lines indicate 95% confidence intervals. (b): Log odds ratio (OR) for severe COVID-19 was calculated by the result of the early second PCR test in patients with a “positive window” and without. OR>0 indicated larger risk in the population that tested positive compared to the population tested negative. Calculated log odds ratios are presented along with gray lines indicating 95% confidence intervals. (c): Same probability analysis for individuals ≥60 years old. (d): Log odds ratio (OR) for severe COVID-19 for individuals ≥60 years old.

### Optimal timing for a second SARS-CoV-2 PCR assessing risk of severe COVID-19 starts at the second day following diagnosis

Our results suggest that a second SARS-CoV-2 test performed in the week following diagnosis could serve as a tool to identify COVID-19 patients at higher risk of developing a severe condition. In order to estimate the best timing to perform the second PCR test, we analyzed test results performed on all adults ≥18 years old every day of the first week and their ability to significantly distinguish between those at a higher risk for severe outcomes (Fig. 5). Data showed that from day 2, and throughout the first week following the first SARS-CoV-2 positive test result, a negative PCR test result was significantly associated with a lower probability for severe disease outcome compared to a positive test result (p<0.05; Supplementary Table 8).

**Fig. 5.**
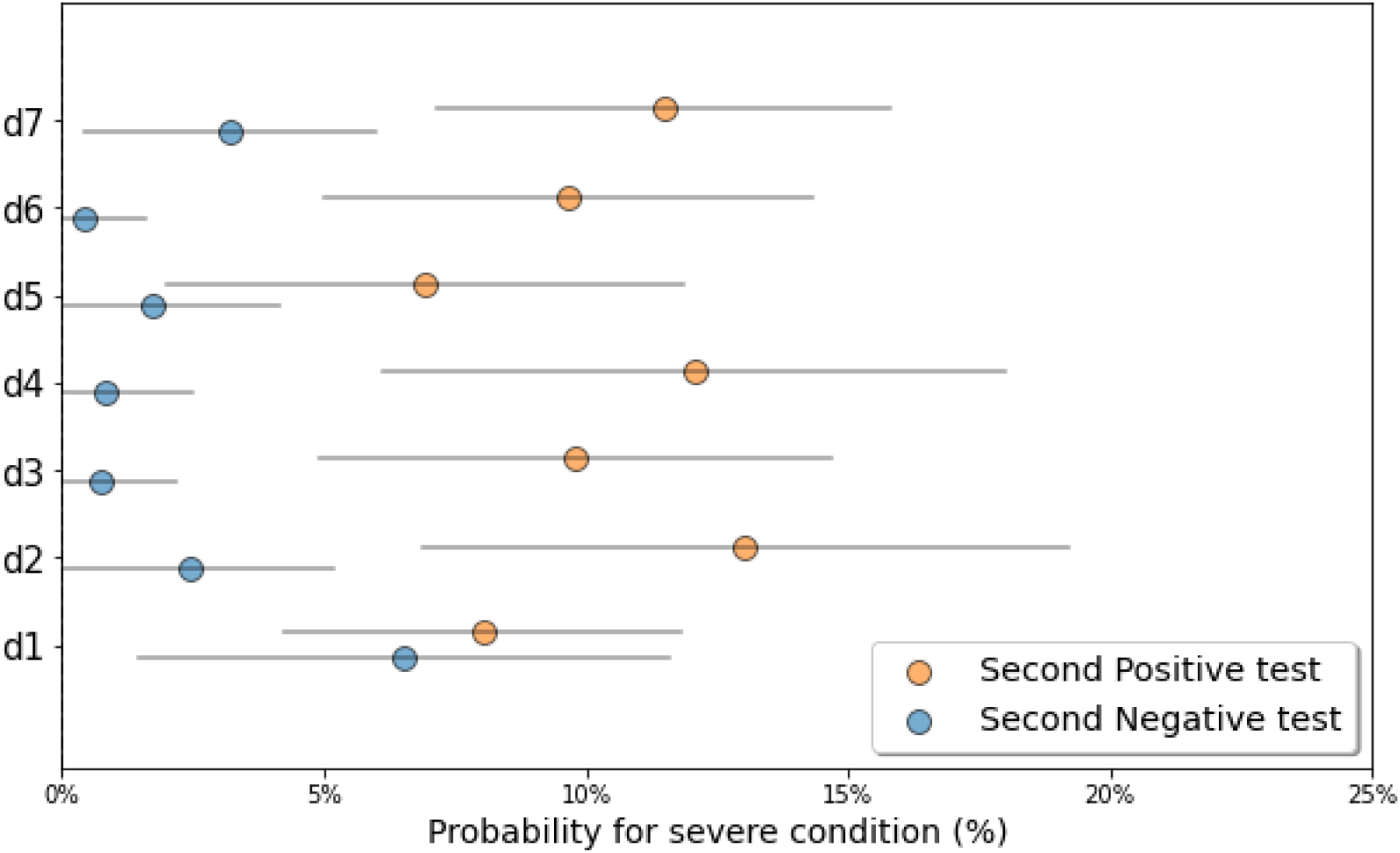
Mapping the best timing to perform the second PCR test to assess risk for disease deterioration following COVID-19 diagnosis. Probability for deterioration to severe COVID-19 in SARS-CoV-2 positive individuals who tested a second time within the first week following their diagnosis. Adult patients were stratified according to the day in which the second test was performed during the first week post diagnosis. Blue circles represent individuals who tested negative and orange circles represent individuals who tested positive. Grey lines indicate 95% confidence intervals.

## Discussion

This study explored the clinical implications of a second PCR testing in COVID-19 infected individuals utilizing EHR data from the second largest HMO in Israel. We analyzed retrospective data starting March 2020, while guidelines for testing were not yet well defined and multiple testing of some individuals were performed in order to understand the clinical course of the SARS CoV-2 infection better. Our analysis demonstrated that a second SARS-CoV-2 PCR test performed 2-7 days from diagnosis, was able to indicate clinical risk of deterioration to severe illness.

Focusing on the population that had a second test within a week of a positive SARS-CoV-2 result enabled us to look whether testing within a short time interval from diagnosis could give indication to future clinical outcomes and assist physicians in disease management of their patients. As time from exposure to symptom onset is 4-5 days and in symptomatic patients disease worsens within 5-10 days from symptom onset ^19,20^, we focused on additional testing within the first week of diagnosis. This is before patients’ clinical course becomes clear and those without severe symptoms are sent home for quarantine, away from medical observation. In agreement with current knowledge regarding risk factors for severe outcomes ^21–23^, our data showed that the probability of deteriorating to severe COVID-19 was age-dependent and was most substantial in adults older than 60 years. While adults under 60 years presented 0.99% probability of deterioration, adults aged 60-79 years and >=80 years presented much higher probabilities, 8.15% and 30.62%, respectively. Despite the basic probability differences, a second SARS-CoV-2 test was significantly associated with subsequent deterioration to severe illness at all ages (OR=4 for 18-59 years, OR=4.15 for 60-79 years and OR=4.88 for adults >80). These observations demonstrate that a second SARS-CoV-2 test may point to patients who will more likely deteriorate to a severe clinical condition.

While hospitalized patients present worse symptoms and have more severe outcomes than patients managing their illness at home, it was important to understand whether the second test result was dependent on the clinical settings at the time of testing. Although there were differences in the probability for severe disease and hospitalized patients were more likely to experience severe outcomes, the second SARS-CoV-2 PCR test was able to identify patients at increased risk for clinical deterioration in both settings. The test result in the second PCR was significantly indicative of severe COVID-19 in patients admitted to the hospital (In adults: OR=2.06 (1.14-3.7), p=0.016; In ≥60 years: OR=2.4 (1.11-5.2), p=0.027) and even more so in patients in the community (In adults: OR=17.56 (2.30-133.93), p=0.006; In ≥60 years: OR=12.92 (1.64-101.2), p=0.015). This demonstrates not only the possible utility of a second PCR test by physicians in both settings, but also the significant value such a test can offer in the community settings to support a more informed clinical decision-making. Previous reports have shown that viral dynamics in patients with mild and severe disease differed and that patients with severe illness had higher viral loads and longer virus shedding period compared to mild cases ^24,25^. This analysis may explain our observations as they report peak viral loads in the second week from disease onset in mild cases while the severe cases show continuous high viral loads at the third and fourth week following disease onset. It has therefore been suggested that detection and quantification of viral RNA levels could aid risk-stratification of hospitalized patients ^26^. Other efforts are continuously directed towards building tools for predicting disease deterioration of hospitalized patients using lungs CT, biomarkers, blood tests values, respiratory values and vital signs ^27–29^. Currently, none of these approaches are focusing on aiding patients stratification while not in the hospital. Using a second PCR test as a measure to assess deterioration risk could assist in risk-stratifying patients and orchestrating care in both settings and with the simple PCR analysis output of positive/negative, without specific viral load values. In patients tested positive for the second time, adjustment of clinical measures may include a closer testing and monitoring schedule and earlier treatment measures to prevent deterioration while a negative test could be the first indication of clinical improvement. Similarly in the community, a second positive test can support physicians’ decisions in prescribing specific home care, maintaining close medical surveillance and regular updates in case hospitalization will be needed in the following days. A negative test within the first week from the first positive PCR test could give a crude indication, even in the older at-risk population, that the patient is at lower-risk to develop a severe disease and guide clinical management accordingly.

An important observation in our data was that even during a patient’s SARS-CoV-2 PCR positive period, when disease is not considered resolved yet, additional tests performed between the first and the last positive PCR tests were indicative of clinical outcome. A second negative PCR result was associated with lower probability to develop severe COVID-19 compared to a positive result and compared to the risk seen in individuals tested positive once and with no subsequent tests. This unique analysis was available to us as vague testing policy in Israel at the beginning of the pandemic resulted in multiple testing of SARS-CoV-2 PCR positive individuals. This created a cohort of patients who were tested longitudinally during the period of viral infection. Negative test results during this period were considered “false negatives” since patients had later evidence of viral RNA, nevertheless, our data demonstrates they correlated with better clinical outcomes. Explanations for this observation may include a lower viral load during the disease timeline, efficient immune response against the virus and possible disease resolution. Previous reports have demonstrated that during an active SARS-CoV-2 infection, viral loads in the host can fluctuate and result in negative test results as they reach levels lower than the tests’ detection limit ^17,30–34^. This study shows that PCR tests can be utilized close to diagnosis for risk assessment and that a negative test result during an active SARS-CoV-2 infection has clinical implications and indicates a milder disease course.

Second PCR testing was able to distinguish between those at higher risk and those at lower risk of deterioration in most days during the first week following the first positive SARS-CoV-2 PCR test. The differences in risk for COVID-19 deterioration were first seen at day two following the positive test and continued through day seven. This observation is highly valuable and could prove useful in clinical settings, when patients are just diagnosed and the physicians need to decide quickly what is the best treatment course, without knowing the patients’ exact disease timeline and what would be the subsequent severity of their illness. An early second PCR test, 2-4 days following the first positive test, may serve as an additional deterioration risk assessment tool for severe COVID-19 before patients start deteriorating and support timely medical care. It may assist treatment decisions such as hospitalization vs home care, frequency of clinical monitoring at the community/home and serve as a preliminary alert for physicians of patients at risk of developing a severe COVID-19. A prospective study is planned to evaluate the effect of this tool on hospitalizations, home-care and treatment approaches in infected COVID-19 individuals.

Our study has several strengths. First, this unique dataset with longitudinal multiple testing following a positive SARS-CoV-2 test result was available as a result of how COVID-19 response in Israel shaped. At the beginning of the pandemic, Israeli citizens could request a COVID-19 test through multiple channels: the national corona focal point, their general practitioner, directly from their HMO or while visiting the ER in the hospitals. As these systems were not synchronized, individuals were able to get tested several times in short intervals close to the date of diagnosis until concrete national guidelines demanded a reference by the physician for COVID-19 test. Additionally, until mid-July 2020, a SARS-CoV-2 positive individual needed 2 consecutive negative tests in order to end isolation, which added data and the ability to detect a “positive window” of individuals with COVID-19. Second, we used HMO data containing all the patients diagnosed with COVID-19 and analyzed results of all tests that were done both in the hospitals and in the community. This makes this study and its conclusions relevant for the general population and valid in the different settings where COVID-19 patients are cared for. Validating the analysis in the different subgroups and observing repeatedly that a second PCR test is indicative of the probability for severe outcomes increased the confidence in these findings.

Our study also has several limitations. First, we were unable to determine what was the reason for the multiple PCR tests performed by the individuals. The additional tests were not done by all SARS-CoV-2 positive patients documented in Maccabi Health Services. Within the week following a positive test we had results of a second PCR test for 10% of the patients 18-59 years old, 14% for ages 60-79 and 32% for patients older than 80. To control for a potential bias, we also analyzed all the patients who tested positive and did not have an early second PCR test. Their risk for severe condition was in the range of those who tested positive/negative the second time within a week from diagnosis. Second, the guidelines for COVID-19 testing in Israel had changed several times during the study period and the inconsistent reasons and frequency for SARS-CoV-2 PCR testing may affect our results.

In conclusion, this study explored a new application for multiple SARS CoV-2 PCR testing. While current multiple testing approaches aim to detect COVID-19 early, prevent transmission, contain it and reduce morbidity and mortality, we suggest that additional PCR testing is used in the clinic as an early, wide-spread complementary tool for risk assessment and subsequent appropriate disease management. This could direct appropriate resources and guide clinical testing and isolation of patients at risk.

## Methods

### Data

Data in this study originated from Maccabi Healthcare Services (MHS) which is the second largest active HMO in Israel. As participation in a medical insurance plan is compulsory in Israel and all citizens must join one of four official Israeli HMOs, there is longitudinal health data on most Israeli citizens. MHS data includes 2.3 million insured citizens starting 1993, with annual attrition rate lower than 1%. The dataset we analyzed here included demographic data, SARS-CoV-2 test results and clinical surveillance in community clinics and hospitals.

### Study outcome

COVID-19 patients were defined as those tested positive in a SARS-CoV-2 polymerase chain reaction (PCR) test obtained from nasopharyngeal swabs. The severe COVID-19 cohort included patients whose disease status deteriorated to severe, admitted to the intensive care unit or died as updated by hospital staff. Initially, the definition varied slightly between hospitals but was commonly dictated by the severity of symptoms in the lower respiratory tract, respiratory distress, pneumonia, use of artificial respiration, shock and system failure. SARS-CoV-2-positive patients that were not reported with a severe disease status, including asymptomatic, mild patients or with unknown status, constitute the cohort of infected COVID-19 patients that did not deteriorate to severe condition.

### Study design and population

We analyzed data of individuals from MHS, who had at least one PCR test for SARS-CoV-2 between March 1, 2020 and August 9, 2020. 239,048 individuals tested for SARS-CoV-2. Overall, 375,558 PCR tests for SARS-CoV-2 were performed during this time period. Patients with COVID-19 infection were identified as those having at least one record of a positive SARS-CoV-2 PCR test in their MHS EHR. Individuals negative to COVID-19 infection were considered as such if all their laboratory tests for SARS-CoV-2 were negative. In total we had 15,822 patients with at least one positive test.

This study focused on a population of confirmed COVID-19 patients who performed additional PCR tests following diagnosis. There were a total of 27,796 additional tests performed on 9,021 SARS-CoV-2-positive individuals. Among them, 1,683 patients had a second PCR test in the first week (2-7 days) following diagnosis. This population was evaluated for its association with COVID-19 severity.

Patients with at least two positive SARS-CoV-2 PCR tests and at least one additional test between them were defined as patients with a COVID-19 “positive window”. We identified 3,135 adults with a “positive window” and used this sub-population to analyze whether an early second negative PCR test could be associated with the patients’ disease severity, even when they still during an active infection period.

Data of COVID-19 patients from the hospitals included hospitalization date, disease severity indication including ICU admission and death. Out of the 15,822 infected patients, 416 (2.63%) had severe outcomes during this study period. We used hospitalization data to define the patients’ outcomes and to determine whether the PCR test was performed during hospitalization or in the community.

### Statistical analysis

For analyzing the association between the SARS-CoV-2 PCR result of the second test taken after diagnosis and severe outcomes, we included the first PCR test result from days 2-7 following diagnosis date (which was determined by the first positive SARS-CoV-2 PCR test). The risk for severe outcome was calculated for sub-groups divided by the occurrence and result of the additional test (positive test, negative test and not tested). In addition, we calculated the odds ratio for severe outcomes for positive and negative PCR test results in the different ages groups. This analysis was performed for several age groups.Odds ratios (OR) were calculated by a logistic regression model using R version 3.5.2.

We further analyzed the connection between the additional test and whether the test was performed during hospitalization or in the community, for that we used the date the patient entered the hospital and the date of the additional test.

Next we analyzed negative tests within a positive window which was defined for patients with at least 2 positive tests, the window range was from the first positive test to the last positive test, we analyzed the results of the tests in this window with relation to the distance in days from the start of the window. For each day we calculate the fraction of negative tests.

To find the relevant days that show significant difference in predicting deterioration to severe outcome we run the analysis for each day taking the test if it was the first one during one week from the first positive test. We used chi square test to compute p-value.

## Data Availability

The data that support the findings of this study originate from Maccabi Health Services. Restrictions apply to the availability of these data and they are therefore not publicly available. Due to restrictions, these data can be accessed only by request to the authors and/or Maccabi Health Services.

## Ethics declarations

The study protocol was approved by Maccabi Health Services’ institutional review board (0024-20-MHS). Informed consent was waived by the IRB, as all identifying details of the participants were removed before the computational analysis.

## Code availability statement

Analysis code is available at git though it is tailored to the data and the fields of the Maccabi Health Services database.

## Supplementary Material

**Supplementary Table 1.** Probability analysis for deterioration to severe condition after receiving test result of a 2nd PCR test done within a week following COVID-19 positive diagnosis

| Age (yrs) | Test results | # of patients | # of patients with severe condition | Probability to develop severe condition | 95% CI |
| --- | --- | --- | --- | --- | --- |
| 18-59 | Positive | 624 | 22 | 3.53% | [2.08%-4.97%] |
|  | Negative | 663 | 6 | 0.90% | [0.18%-1.63%] |
|  | Not tested | 12191 | 106 | 0.87% | [0.70%-1.03%] |
| 60-79 | Positive | 158 | 31 | 19.62% | [13.43%-25.81%] |
|  | Negative | 108 | 6 | 5.56% | [1.24%-9.88%] |
|  | Not tested | 1673 | 121 | 7.23% | [5.99%-8.47%] |
| ≥80 | Positive | 91 | 38 | 41.76% | [31.63%-51.89%] |
|  | Negative | 39 | 5 | 12.82% | [2.33%-23.31%] |
|  | Not tested | 275 | 81 | 29.45% | [24.07%-34.84%] |
| All | Positive | 873 | 91 | 10.42% | [8.40%-12.45%] |
|  | Negative | 810 | 17 | 2.10% | [1.11%-3.09%] |
|  | Not tested | 14139 | 308 | 2.18% | [1.94%-2.42%] |

**Supplementary Table 2.** Odds ratio (OR) analysis of a second positive SARS-CoV-2 PCR test result within a week of COVID-19 diagnosis and deterioration to severe COVID-19.

| Age (yrs) | OR | 95%CI | p-value |
| --- | --- | --- | --- |
| 18-59 | 4.00 | [1.61-9.94] | 0.0028 |
| 60-79 | 4.15 | [1.67-10.33] | 0.0022 |
| ≥80 | 4.88 | [1.75-13.62] | 0.0025 |

**Supplementary Table 3.**

| Age (yrs) | Test within a week of diagnosis | # of patients | # of patients with severe condition | Probability to develop severe condition | 95% CI |
| --- | --- | --- | --- | --- | --- |
| Admitted to hospital $\geq 18$ yrs | Positive test | 304 | 77 | 25.33% | [20.44%-30.22%] |
|  | Negative test | 113 | 16 | 14.16% | [7.73%-20.59%] |
|  | Performed a second test | 417 | 93 | 22.30% | [18.31%-26.30%] |
| In the community $\geq 18$ yrs | Positive test | 569 | 14 | 2.46% | [1.19%-3.73%] |
|  | Negative test | 697 | 1 | 0.14% | [0.00%-0.42%] |
|  | Performed a second test | 1266 | 15 | 1.18% | [0.59%-1.78%] |
| $\geq 18$ yrs | Positive test | 873 | 91 | 10.42% | [8.40%-12.45%] |
|  | Negative test | 810 | 17 | 2.10% | [1.11%-3.09%] |
|  | Performed a second test | 1683 | 108 | 6.42% | [5.25%-7.59%] |
| Admitted to hospital $\geq 60$ yrs | Positive test | 145 | 57 | 39.31% | [31.36%-47.26%] |
|  | Negative test | 47 | 10 | 21.28% | [9.58%-32.98%] |
|  | Performed a second test | 192 | 67 | 34.90% | [29.09%-40.70%] |
| In the community $\geq 60$ yrs | Positive test | 104 | 12 | 11.54% | [5.40%-17.68%] |
|  | Negative test | 100 | 1 | 1.00% | [0.00%-2.95%] |
|  | Performed a second test | 204 | 13 | 6.37% | [3.12%-9.62%] |
| $\geq 60$ yrs | Positive test | 249 | 69 | 27.71% | [22.15%-33.27%] |
|  | Negative test | 147 | 11 | 7.48% | [3.23%-11.74%] |
|  | Performed a second test | 396 | 80 | 20.20% | [16.60%-23.81%] |

**Supplementary Table 4.**

| Age (yrs) | Odds ratio | 95%CI | p-value |
| --- | --- | --- | --- |
| Admitted to hospital $\geq 18$ yrs | 2.06 | [1.14-3.70] | 0.0164 |
| In the community $\geq 18$ yrs | 17.56 | [2.30-133.93] | 0.0057 |
| All adults $\geq 18$ yrs | 5.43 | [3.20-9.20] | <0.0001 |
| Admitted to hospital $\geq 60$ yrs | 2.40 | [1.11-5.20] | 0.027 |
| In the community $\geq 60$ yrs | 12.92 | [1.64-101.2] | 0.015 |
| All adults $\geq 60$ yrs | 4.74 | [2.42-9.30] | <0.0001 |

**Supplementary Table 5.**

| Day from diagnosis | # of positive test results | # of negative test result | Total # of tests | Probability for a negative test |
| --- | --- | --- | --- | --- |
| 1 | 112 | 5 | 117 | 4.27% |
| 2 | 77 | 8 | 85 | 9.41% |
| 3 | 104 | 11 | 115 | 9.57% |
| 4 | 99 | 20 | 119 | 16.81% |
| 5 | 89 | 30 | 119 | 25.21% |
| 6 | 128 | 33 | 161 | 20.50% |
| 7 | 185 | 42 | 227 | 18.50% |
| 8 | 164 | 52 | 216 | 24.07% |
| 9 | 171 | 73 | 244 | 29.92% |
| 10 | 206 | 84 | 290 | 28.97% |
| 11 | 258 | 115 | 373 | 30.83% |
| 12 | 604 | 307 | 911 | 33.70% |
| 13 | 390 | 192 | 582 | 32.99% |
| 14 | 251 | 132 | 383 | 34.46% |
| 15 | 206 | 129 | 335 | 38.51% |
| 16 | 179 | 92 | 271 | 33.95% |
| 17 | 177 | 84 | 261 | 32.18% |
| 18 | 146 | 83 | 229 | 36.24% |
| 19 | 201 | 121 | 322 | 37.58% |
| 20 | 165 | 105 | 270 | 38.89% |
| 21 | 112 | 81 | 193 | 41.97% |

**Supplementary Table 6.**

|  | Test result | # of patients | # of patients with severe condition | Probability to develop severe condition | 95% CI |
| --- | --- | --- | --- | --- | --- |
| Positive diagnosis >=18 | Positive | 873 | 91 | 10.42% | [8.40%-12.45%] |
|  | Negative | 810 | 17 | 2.10% | [1.11%-3.09%] |
|  | No test | 14139 | 308 | 2.18% | [1.94%-2.42%] |
| Positive window >=18 | Positive | 563 | 59 | 10.48% | [7.95%-13.01%] |
|  | Negative | 124 | 4 | 3.23% | [1.20%-6.34%] |
|  | No test | 4944 | 172 | 3.48% | [2.97%-3.99%] |
| No Positive window >=18 | Positive | 310 | 32 | 10.32% | [6.94%-13.71%] |
|  | Negative | 686 | 13 | 1.90% | [0.87%-2.92%] |
|  | No test | 9195 | 136 | 1.48% | [1.23%-1.73%] |
| Positive diagnosis >=60 | Positive | 249 | 69 | 27.71% | [22.15%-33.27%] |
|  | Negative | 147 | 11 | 7.48% | [3.23%-11.74%] |
|  | No test | 1948 | 202 | 10.37% | [9.02%-11.72%] |
| Positive window >=60 | Positive | 158 | 42 | 26.58% | [19.69%-33.47%] |
|  | Negative | 24 | 2 | 8.33% | [0.00%-19.39%] |
|  | No test | 876 | 113 | 12.90% | [10.68%-15.12%] |
| No Positive window >=60 | Positive | 91 | 27 | 29.67% | [21.43%-37.91%] |
|  | Negative | 123 | 9 | 7.32% | [2.87%-11.76%] |
|  | No test | 1072 | 89 | 8.30% | [6.72%-9.89%] |

**Supplementary Table 7.**

| group | pos | severe | Neg | severe | Odds ratio | 95%CI | p-value |
| --- | --- | --- | --- | --- | --- | --- | --- |
| Positive window >=18 | 563 | 59 | 124 | 4 | 3.512 | [1.251-9.857] | 0.017 |
| No Positive window >=18 | 310 | 32 | 686 | 13 | 5.447 | [2.820-10.523] | <0.0001 |
| Positive diagnosis >=18 | 873 | 91 | 810 | 17 | 4.967 | [2.933-8.410] | <0.0001 |
| Positive window >=60 | 158 | 42 | 24 | 2 | 3.983 | [0.898-17.671] | 0.0691 |
| No Positive window >=60 | 91 | 27 | 123 | 9 | 5.344 | [2.376-12.062] | 0.0001 |
| Positive diagnosis >=60 | 249 | 69 | 147 | 11 | 4.739 | [2.415-9.301] | <0.0001 |

**Supplementary Table 8.**

| Time following diagnosis (d) | Test result | # of patients | # of patients with severe condition | Probability to develop severe condition | p-value |
| --- | --- | --- | --- | --- | --- |
| **d1** | Positive | 199 | 16 | 8.04% |  |
|  | Negative | 92 | 6 | 6.52% | 0.42 |
| **d2** | Positive | 115 | 15 | 13.04% |  |
|  | Negative | 123 | 3 | 2.44% | 0.002 |
| **d3** | Positive | 143 | 14 | 9.79% |  |
|  | Negative | 134 | 1 | 0.75% | 0.001 |
| **d4** | Positive | 116 | 14 | 12.07% |  |
|  | Negative | 119 | 1 | 0.84% | <0.001 |
| **d5** | Positive | 101 | 7 | 6.93% |  |
|  | Negative | 114 | 2 | 1.75% | 0.06 |
| **d6** | Positive | 155 | 15 | 9.68% |  |
|  | Negative | 116 | 0 | 0.00% | <0.001 |
| **d7** | Positive | 209 | 24 | 11.48% |  |
|  | Negative | 156 | 5 | 3.21% | 0.003 |

## Acknowledgments

We thank the following for their contributions to our efforts:

## Author Contribution

These authors contributed equally: Barak Mizrahi, Maytal Bivas-Benita. B.M. and M.B.B. conceived the project, designed and conducted the analyses, interpreted the results and wrote the manuscript. N.K., P.A. and C.Y. directed the project, designed the analysis and interpreted the results. Y.K and E.R. designed and conducted the analyses. S.H.A. and

G.C. directed the project, provided the data and interpreted the results.

## Corresponding author

Barak Mizrahi, KI Research Institute, 11 Hazayit st, Kfar Malal, Israel.

